# Myo-inositol as a spectroscopic marker of neuroinflammation in frontotemporal lobar degeneration

**DOI:** 10.64898/2026.09.18.26363386

**Authors:** Sean YW Tan, Michelle Naessens, P Simon Jones, Emily G Todd, Rebecca S. Williams, Christopher T. Rodgers, Laura E. Hughes, Noah L Shapiro, Julia Goddard, Meira van Schaik, Amanda Heslegrave, Henrik Zetterberg, Alexander G Murley, Maura Malpetti, James B Rowe

## Abstract

**Background and Objectives:** Cost-effective and mechanistically-informative biomarkers are needed to advance novel therapeutics for dementia. We focus on neuroinflammation in disorders associated with frontotemporal lobar degeneration (FTLD). Commonly used assays include positron emission tomography (PET) imaging and peripheral blood markers. However, PET is limited by radioactivity, which impedes repeated measurements, while peripheral markers are indirect measures of neuropathology. This study investigates the potential of proton magnetic resonance spectroscopy (MRS) - a non-invasive technique that can be deployed alongside structural imaging sequences - to quantify myo-inositol, an inflammatory metabolite found in astroglial and microglial cells. We test the following hypotheses (1) myo-inositol is elevated in brain regions affected by FTLD disorders; (2) myo-inositol in affected brain regions correlates with peripheral immune markers; and (3) myo-inositol in affected brain regions correlates with cognitive decline.

**Methods:** 54 people with FTLD-associated disorders (29 behavioural variant frontotemporal dementia, 25 progressive supranuclear palsy) and 73 age/sex-matched controls underwent single-voxel 7 Tesla MRS. 2cm isotropic voxels were placed over the right inferior frontal gyrus (R IFG, a disease-affected region of interest), and right occipital lobe (R OCC, a control region). Serum inflammatory markers were measured using nucleic acid-linked immuno-sandwich assays (NULISA). Cognition was assessed with the revised Addenbrooke’s Cognitive Examination (ACE-R). Mixed-effects and general linear models were used to evaluate diagnostic differences in regional myo-inositol and its association with cognition. Partial least squares regression identified two serum inflammatory components that covaried with myo-inositol; these components were subsequently tested in confounder-adjusted second-level linear models.

**Results:** Myo-inositol was elevated in the R IFG (β = 0.20, t = 5.10, p = 8.8 × 10^-7) but not in the R OCC (β = 0.05, t = 1.36, p = 0.18), in people with FTLD versus controls. In FTLD, myo-inositol in the R IFG was positively associated with a peripheral inflammatory profile characterised by IL-6, IL-1β, GFAP, CSF2, CRP, and CCL22 (β = 0.39, t = 5.12, p = 3.60 × 10^-6). R IFG myo-inositol was negatively associated with ACE-R performance in patients (β = −44.59, t = −4.12, p = 0.00016).

**Conclusion:** MRS-derived myo-inositol is a promising neuroimaging biomarker of inflammation in FTLD. Elevated levels in disease-affected regions were associated with peripheral markers of inflammation (reflecting astrocytosis and microgliosis) and with cognitive impairment.

## Introduction

The syndromes associated with frontotemporal lobar degeneration (FTLD) encompass a pathologically and clinically heterogeneous spectrum, including the behavioural variant frontotemporal dementia (bvFTD)^1^ and progressive supranuclear palsy (PSP)^2^. Although these syndromes vary in their presentation and underlying molecular pathology, they share critical aspects of neuropathology and the pressing need for new therapeutic strategies. In both bvFTD and PSP, there is evidence of neuroinflammation as a common underlying pathophysiological process^3^ in pre-clinical studies^4^, genetic associations^5,6^, neuropathology^7,8^, and positron emission tomography (PET)^9–11^. Neuroinflammation includes astroglial and microglial activation, and increased secretion of cytokines^3^. Neuroinflammation can be quantified *in vivo* using ligands for the 18 kDa translocator protein (TSPO PET), such as ¹¹C-PK11195. These serve as proxies for microglial burden^12^ and are increased in each of the syndromes associated with FTLD^9–11,13^. The neuroinflammation co-localises with other markers of molecular pathology^10,11^ and predicts both cognitive and clinical decline^11,14^, and survival^15^.

Immunotherapeutic strategies therefore have the potential to attenuate progression in FTLD-related syndromes. However, therapeutic development requires scalable and informative biomarkers of neuroinflammation. Blood biomarkers including cytokine profiles and cellular immunophenotypes are abnormal in all types of FTLD, and correlate with severity and progression^16–18^. However, blood biomarkers are not direct or regionally specific markers of neuroinflammation. In contrast, brain imaging markers of inflammation can quantify and directly localise neuroinflammation. For example, TSPO PET measures of microglial activation are increased in disease-specific regional patterns and predict survival in FTD and PSP^14,15^. However, PET requires specialist facilities and radiation exposure, reducing its suitability for repeated use in clinical trials or for stratification. An alternative imaging modality to assay neuropathological processes is magnetic resonance spectroscopy (MRS).

MRS is a non-invasive method that quantifies the chemical composition of brain tissue^19^. It leverages standard magnetic resonance imaging technology and can be integrated as an additional sequence during routine structural imaging, without contrast agents. One of the metabolites quantified by proton-MRS is myo-inositol (mI), a putative marker of astrocytic and microglial activation^20–24^. Elevated mI has been reported in symptomatic FTLD patients^24–26^ and pre-symptomatic mutation carriers^27,28^. Elevated myo-inositol has also been reported in other neuroinflammatory conditions, from COVID-19^29^ and multiple sclerosis^30^, and correlates with their disease severity. Myo-inositol derived from spectroscopy may therefore constitute a candidate in vivo neuroimaging biomarker of inflammation.

Uncertainties remain over the utility of myo-inositol as a marker of neuroinflammation. First, the elevation of myo-inositol in FTLD is reproduced in some studies but not all^31,32^. Second, if myo-inositol reflects underlying neuroinflammation, elevated levels should correlate with other markers, such as peripheral immune markers, similar to the correlations between TSPO PET and serum cytokines in FTLD disorders^17^. This remains to be established. Third, if myo-inositol is pathophysiologically relevant, it should correlate with disease severity, as for TSPO PET^14^.

This study assessed the utility of myo-inositol as a neuroinflammatory biomarker by testing three hypotheses: (1) myo-inositol is selectively elevated in the prefrontal cortex in FTLD disorders; (2) prefrontal myo-inositol correlates with peripheral immune markers; and (3) prefrontal myo-inositol correlates inversely with cognitive function.

## Materials and Methods

### Participants

58 patient-participants were recruited from the Cambridge Centre for Frontotemporal Dementia and the Cambridge Centre for Parkinson-Plus, meeting clinical diagnostic criteria for probable PSP Richardson’s syndrome^2^ or bvFTD^1^. 74 age/sex-matched controls with no history of significant neurological or psychiatric illness were recruited either through Joint Dementia Research or the Cognition and Brain Sciences Unit healthy volunteer panel. Cognition was assessed using the revised Addenbrooke’s Cognitive Examination (ACE-R)^33^. Four patients and one control were excluded due to poor imaging quality, described below.

### Structural imaging and MRS acquisition

Participants underwent scanning with a 7Tesla MAGNETOM Terra scanner (Siemens Healthineers) with a 32-channel receiver and single channel transmit head coil (Nova Medical). A T1-weighted MP2RAGE structural sequence [repetition time = 4300 ms, echo time = 1.99 ms, resolution = 99 ms, bandwidth = 250 Hz/px, voxel size = 0.75 mm3, field of view = 240 × 240 × 157 mm, acceleration factor (A ≫ P) = 3, flip-angle = 5/6° and inversion times = 840/2370 ms] was acquired for voxel placement and CSF partial volume correction. Single voxel proton magnetic resonance spectroscopy (2 cm x 2cm x 2 cm) was acquired from the right inferior frontal gyrus (RIFG) as the disease region of interest, and the right primary visual cortex (ROCC) as a control region. The RIFG was selected *a priori* because spectroscopic abnormalities and disrupted functional connectivity in this region have been associated with cognitive impairment and behavioural dysregulation in frontotemporal lobar degeneration syndromes^32,34–36^. Voxels were placed manually by trained operators using anatomical landmarks. Spectra were acquired using a short-echo semi-LASER sequence (64 repetitions, TR/TE = 5000/28 ms)^37,38^ with FASTESTMAP shimmin^39^ and water-peak flip angle and VAPOR water suppression^40^.

### MRS processing

The 64 individual repetitions were saved separately then preprocessed using FSL MRS version 2.1.13^41^. They were converted into a NifTI-MRS format using spec2nii version 0.7.1^42^ and pre-processed using a standard *fsl_mrs_preproc* pipeline, including coil combination using a whitened singular value decomposition approach, eddy-current correction based on the unsuppressed water reference, and frequency and phase alignment of individual transients using spectral registration. Transients identified as outliers (>2.5 SD from the mean) were excluded prior to averaging. The remaining transients were averaged to produce a single spectrum, and residual water signal was removed using the Hankel–Lanczos singular value decomposition algorithm. Spectral fitting was then performed using the FSL-MRS fitting framework, which employs linear-combination modelling of metabolite basis spectra with parameter estimation using a Markov chain Monte Carlo (MCMC) algorithm^41^. Metabolite resonances between 0.2 and 4.2 ppm were fitted using a simulated basis set that incorporated experimentally acquired macromolecule spectra^36^.

Metabolite concentrations were quantified relative to the unsuppressed water signal and are reported in units of molality (mmol/kg). Metabolite concentrations were log2 transformed to normalise their distributions and to match the logarithmic space of blood-based biomarkers in subsequent analyses. To account for differences in tissue composition within the spectroscopy voxel, partial-volume estimates of grey matter (GM), white matter (WM), and cerebrospinal fluid (CSF) were obtained using the *svs_segment* tool within FSL-MRS. This performs tissue segmentation of the structural T1-weighted image using the FSL *fsl_anat*/FAST segmentation pipeline and calculates the fractional tissue volumes corresponding to the MRS voxel. These tissue fractions were incorporated into the water-referencing model to correct metabolite estimates for CSF contamination.

Quality control assessments included (1) ensuring the full width half maximum (FWHM) of the unsuppressed water peak being < 16 Hz. (2) Cramer Ro Lower Bound (CRLB) < 20 for metabolites of interest (3) inspection of voxel placements and metabolite spectra (4) transients were adequate or better, according to the published MRS Expert Consensus guidelines for the brain^43^. Voxel placements from all participants were co-registered and overlaid using a T1 template in MNI space (Figure 1) (4) Comparison of signal to noise ratio (SNR), FWHM and CRLB for myo-inositol between groups (Supplementary Tables 1 and 2).

**Figure 1.**
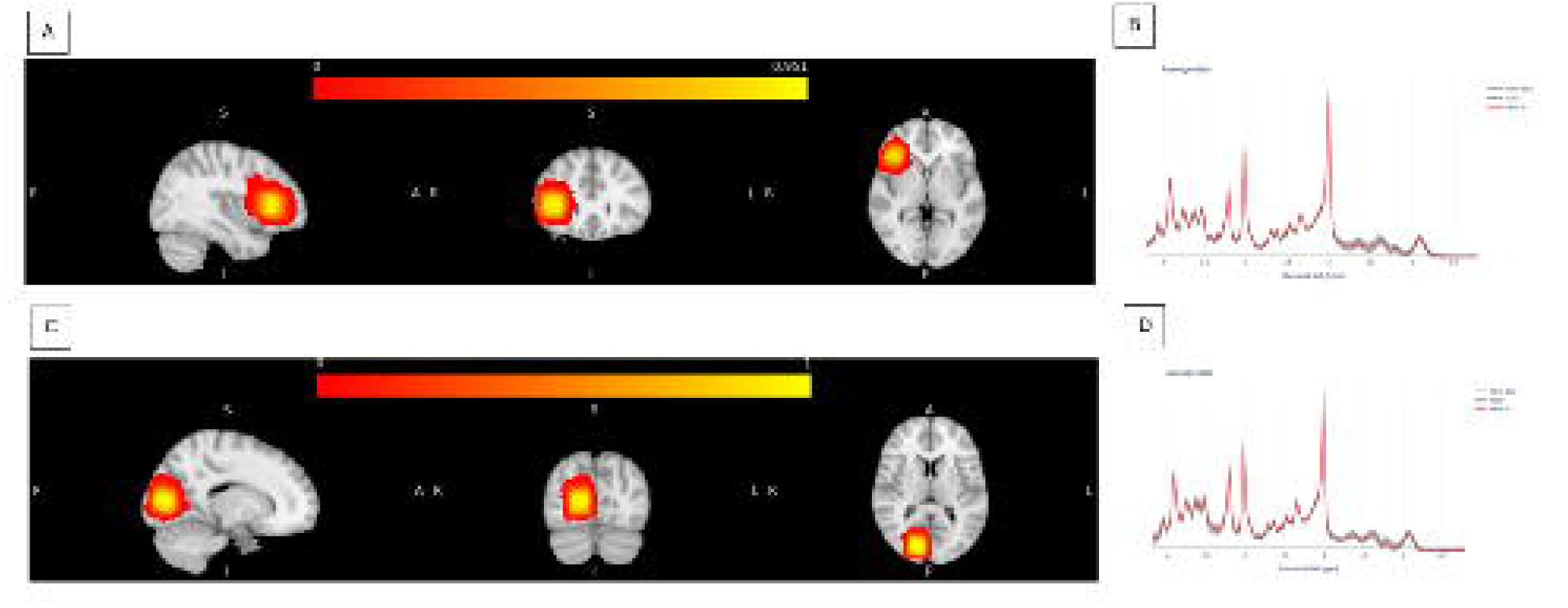
Voxels from all participants are overlaid in MNI space to show the locations of the right inferior frontal gyrus (A) and right occipital lobe (C) voxels. The scales above the iinages indicate the fraction of overlap (0 = no overlap, 1 = complete overlap). Su1nmary spectra from both regions are also shown (B, D).

As a comparator metabolite, we assessed N-acetylaspartate (NAA) using the same modelling approach, to confirm that increases in myo-inositol were not simply an artifact from CSF correction. As NAA would be expected to be static or reduced in the disease^25,32,44^, the presence of such a decrease would support the view that a rise in myo-inositol was not an artefactual inflation from volume-correction.

### Blood sampling and processing

Plasma samples were obtained and processed as described previously^16^. Briefly, blood samples were collected by venepuncture in ethylenediaminetetraacetic acid tubes. They were centrifugated to separate plasma, aliquoted and stored at −70◦C. Plasma samples were analysed using nucleic acid-linked immuno-sandwich assays CNS120 panel (Alamar Biosciences, NULISASeq)^45^. The 52 markers of inflammation within the CNS panel were selected for further analysis (Supplementary Table 4 for the list of inflammatory markers used in subsequent analysis). Data were normalized using internal and inter-plate controls, then rescaled by multiplication with a factor of 10^4. Next, +1 was added to all values before log2 transformation. These values are defined as NULISA Protein Quantification (NPQ) units, on the logarithmic scale.

### Statistical analysis

This was a retrospective cross-sectional study, with statistical analysis using R version 4.5.2. Differences in categorical variables were assessed using chi-square, whereas t-tests, ANOVAs, and post-hoc Tukey’s Honest Significant Difference tests were used for continuous variables. Statistical significance was set at p < 0.05, with Tukey’s p-value adjustment applied where appropriate. Further inferential analyses were performed using general linear models and linear mixed-effects models (using the *lm and lmer* functions), with the corresponding equations described below. Assessment of influence diagnostics revealed no outliers unduly influenced model results (all Cook’s distance values < 0.5). Effects of interest were extracted for further analysis using the *emmeans* (estimated marginal means) and *emtrends* (slope parameters) functions. Missing data for ACE-R scores and years of education were present in five participants and were addressed using multiple imputation with the *mice* package.

Analysis first examined bvFTD and PSP as a combined group (“FTLD”) in comparison with controls and followed by separate 3-group analysis (bvFTD, PSP, Control). The sections below describe the analysis of (1) regional differences in MRS derived myo-inositol (2) linking myo-inositol in the RIFG to peripheral blood and (3) linking myo-inositol in the RIFG to cognitive function.

### MRS analysis

127 participants had usable MRS data from the RIFG. Among these, 121 participants also had usable MRS data from the right OCC region. This smaller sample was used to assess inter-regional differences in myo-inositol and its interactions with diagnostic group. Analyses of myo-inositol differences in the RIFG alone were conducted using the full sample of 127 participants. The mixed effect model used to assess for inter-regional differences and interactions with diagnosis used the following model to account for covariates and repeated measures:

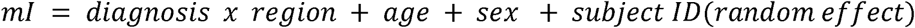

The same type of mixed-effects model was also used to assess differences in brain tissue fraction (1–CSF) within the voxels, to demonstrate that the RIFG was affected in the disease groups, with brain tissue fraction as the dependent variable (right hand side of equation).

Assessing myo-inositol and NAA changes only within the RIFG used the linear model below:

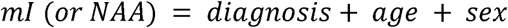

### MRS to blood analysis

65 participants (29 controls, 18 bvFTD, 18 PSP) had NULISA assays that passed quality checks within one year of imaging. The mean ± standard deviation of the time difference between blood sampling and imaging was −0.08 ± 0.29 years for controls and −0.14 ± 0.32 years for FTLD. Time differences did not differ between groups (t (61.75) = 0.91, p = 0.365). The 52 inflammatory blood tests were z scored against means of the entire cohort and underwent batch effect correction with COMBAT^46^. Myo-inositol values were also z-scored after the log2 transformation.

Partial least squares regression (PLS) was used to identify latent components of serum markers that best covaried with myo-inositol, using the *pls* package with the *plsr* function and leave-one-out cross-validation. The number of PLS components selected for further analysis (n=2) was determined using non-parametric bootstrapping with the *bootPLS* package via the *nbcomp.bootplsR* function. Fifty runs of 500 bootstrap replicates each were conducted to ensure consistency in the number of recommended components across runs. The first two components explained 53.16% of the variance in myo-inositol (component 1: 30.73%; component 2: 22.43%), and these two components were examined in subsequent analyses.

To assess the relationship between these PLS components and myo-inositol, and how this differed between diagnostic groups while accounting for covariates, a second-level model was employed^47^. The component scores were placed into a linear model with the following specification:

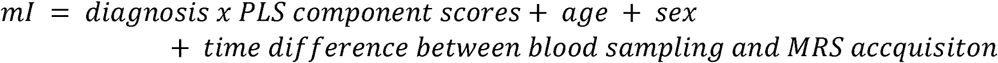

### MRS to cognition

The relationship between myo-inositol in the RIFG and cognition was examined in patients. ACE-R assessments closest to the time of imaging were used (max 1 year). 3/54 patients were excluded from analysis. Across all patients, the mean ± standard deviation of the time difference between ACE-R assessment and imaging was 0.01 ± 0.22 years, with all assessments occurring within half a year of imaging (range = −0.46 to 0.52 years). There was no significant difference between bvFTD and PSP patients (t (43.9) = −0.54, p = 0.59).

To assess the relationship between ACE-R score and myo-inositol, the following linear model was used for the patient group as a whole:

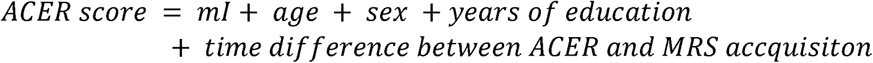

When bvFTD and PSP subgroups were examined, the same model was used with the addition of an interaction term between diagnosis and myo-inositol (diagnosis × mI). The mean ± standard deviation of years of education for the respective subgroups were 12.4 ± 3.36 years for bvFTD and 12.7 ± 2.69 years for PSP. There was no significant difference between groups (t (47.4) = −0.41, p = 0.68).

### Standard Protocol Approvals, Registrations and Patient Consents

Participants with mental capacity gave their written informed consent to take part in the study according to the Declaration of Helsinki. For those who lacked capacity, their participation followed the personal consultee process in accordance with the UK law. The research protocols were approved by the National Research Ethics Service’s East of England Cambridge Central Committee (REC references: 16/EE/0351,16/EE/0084).

## Results

### Cohort characteristics

Table 1 summarises the demographic characteristics of the sample used in this analysis. Patients and controls did not differ in age or sex. FTLD patients showed impairment on the ACE-R compared to controls, with the greatest deficit observed in the bvFTD subgroup.

**Table 1.**
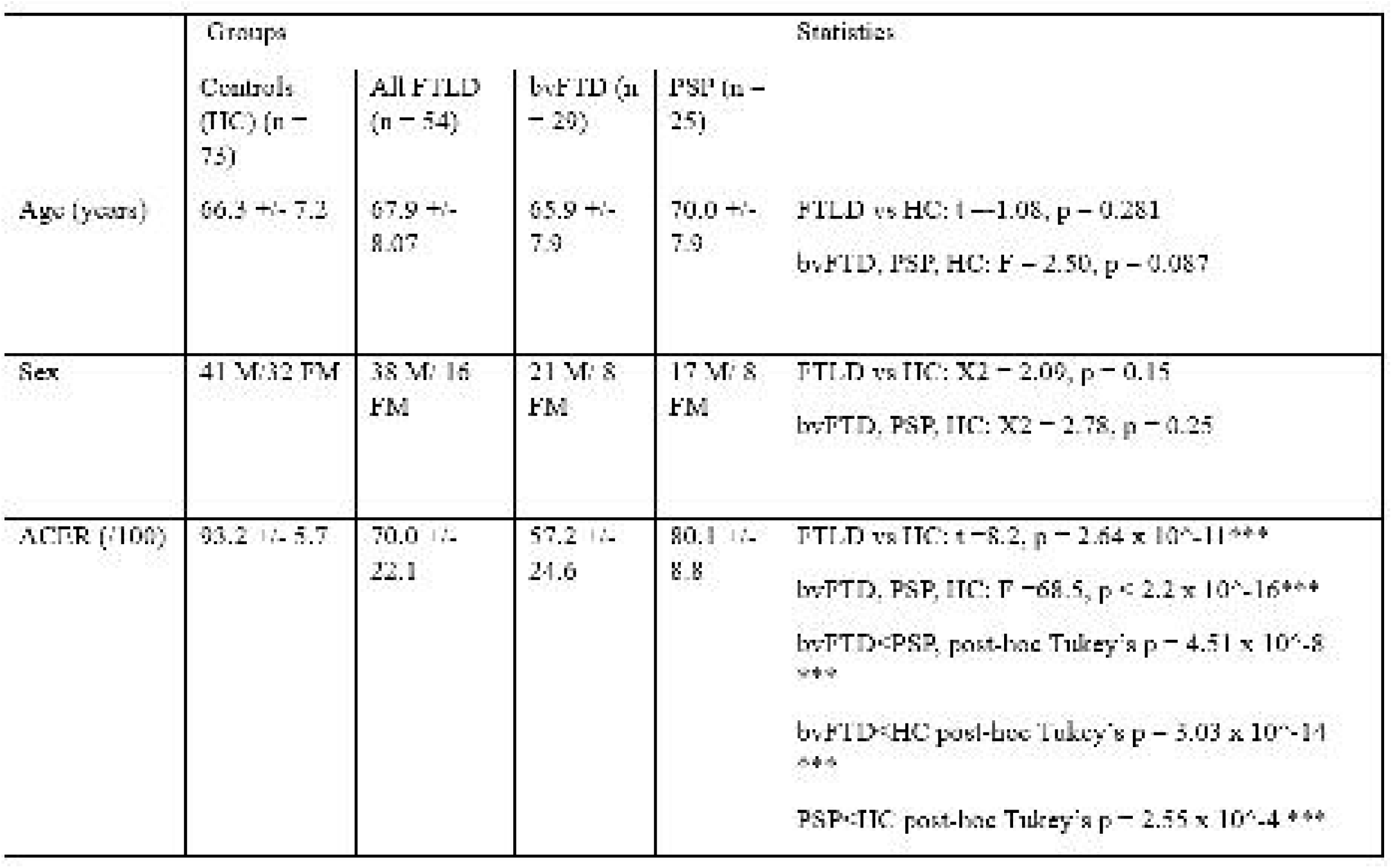
Demographic characteristics of the sample. Values are presented as mean +/. standard deviation. Statistics are derived from chi-square, t-tests, ANOVAs and post-hoc Tukey’s honest significant difference tests. *** p < 0.001

### Myo-inositol is elevated in the prefrontal cortex in FTLD disorders

The fraction of brain tissue is reduced in the RIFG in FTLD disorders (β = −0.09, standard error (SE) = 0.009, t = −9.53, p < 2.2 × 10^-16) compared to controls. While a difference was also observed in the ROCC region (β = −0.02, SE = 0.009, t = −2.08, p = 0.04), the magnitude of the decrease was smaller than in the RIFG (β = −0.07, SE = 0.01, t = −5.45, p = 2.91 × 10^-7; Supplementary Fig 1). Subgroup analysis (Supplementary Fig 2) revealed that atrophy within the RIFG was greatest in the bvFTD subgroup compared to controls (β = −0.13, SE = 0.01, t = −12., p < 2.2 × 10^-16) and PSP (β = −0.09, SE = 0.01, t = −6.81, p = 8.13 × 10^-11). PSP showed an intermediate degree of volume loss, relative to controls (β = −0.04, SE = 0.01, t = −4.12, p = 0.0002).

Myo-inositol was elevated in the RIFG in FTLD patients compared to controls (β = 0.20, SE = 0.04, t = 5.10, p = 8.8 × 10^-7), whereas no significant difference was observed in the ROCC region (β = 0.05, SE = 0.04, t = 1.36, p = 0.18) (Figure 2A). The elevation between groups in the R IFG differed significantly from the difference in groups in the ROCC controls (β = 0.25, SE = 0.04, t = 6.99, p = 1.74 × 10^-10). Subgroup analysis showed that the greatest myo-inositol increase within the RIFG was in the bvFTD cohort (Figure 2B) with the following effect sizes noted for bvFTD compared to controls (β = 0.29, SE = 0.05, t = 5.98, p = 1.3 × 10^-8) and PSP (β = 0.19, SE = 0.06, t = 3.08, p = 0.0068). PSP participants showed an intermediate degree of elevation compared to controls that did not achieve statistical significance (β = 0.11, SE = 0.05, t = 2.19, p = 0.08).

**Figure 2.**
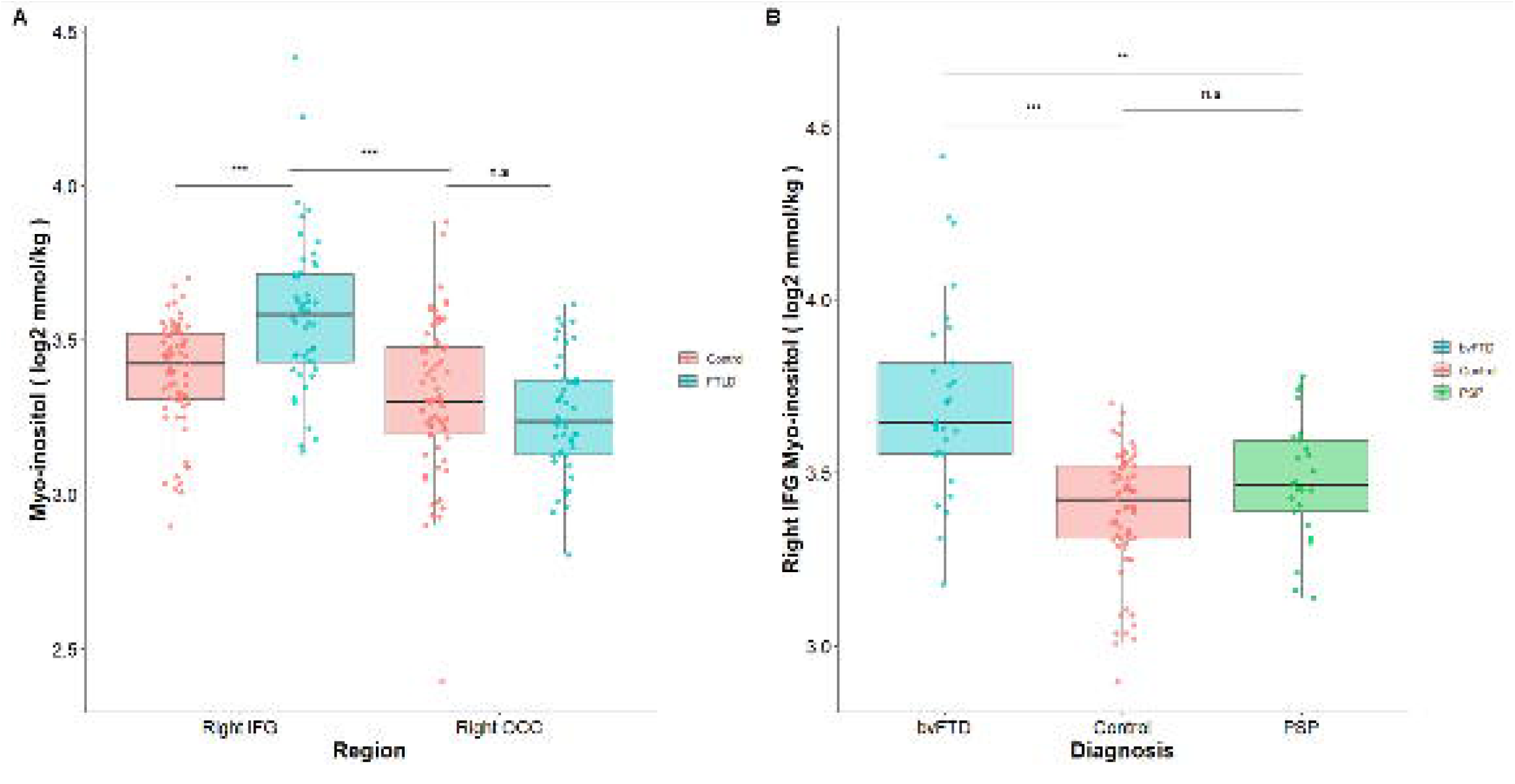
Boxplots showing myo-inositol levels across brain regions (right inferior frontal gyrns and right occipital lobe voxels) in control and FTLD groups (A). Boxplots showing myo-inositol levels within the right inferior frontal gyms voxel **in** control, bvFTD and PSP pru·ticipants (B). *** p < 0.001; ** p < 0.01; n.s = not significant

Analysis of the RIFG in the full sample of 127 participants replicated these findings, demonstrating elevated mI levels in FTLD patients (β = 0.22, SE = 0.04, t = 6.04, p = 1.71 × 10^-8; Supplementary Figure 3). Differences in subgroups were still present after the adjustment for confounds (F (2, 122) = 27.39, p = 1.50 × 10^-10), with the highest difference observed in bvFTD (Supplementary Figure 4, Supplementary Table 5).

NAA was reduced in FTLD patients (β = −0.20, SE = 0.03, t = −6.40, p = 2.92 × 10^-9) (Supplementary Figure 5). Subgroup differences remained present after adjustment for confounders (F (2, 122) = 53.20, p < 2 × 10^-16; see also Supplementary Figure 6, Supplementary Table 6).

### Associations between myo-inositol and peripheral immune markers in patients

The first two latent components derived from the PLS analysis were extracted and their biomarker loadings are shown in Figure 3. The first latent component was characterised by broadly positive loadings across most inflammatory markers, whereas the second latent component showed a selective loading profile, with positive loadings for Interleukin-6 (IL-6), interleukin-1 beta (IL-1β), glial fibrillary acidic protein (GFAP), colony-stimulating factor 2 (CSF2), C-reactive protein (CRP), and C-C motif chemokine ligand 22 (CCL22). FTLD patients showed higher scores on PLS component 1 relative to controls t (69.10) = 3.82, p = 0.0004. However, no significant association was found between PLS component 1 and mI levels for FTLD patients (β = 0.10, SE = 0.06, t = 1.63, p = 0.11).

**Figure 3.**
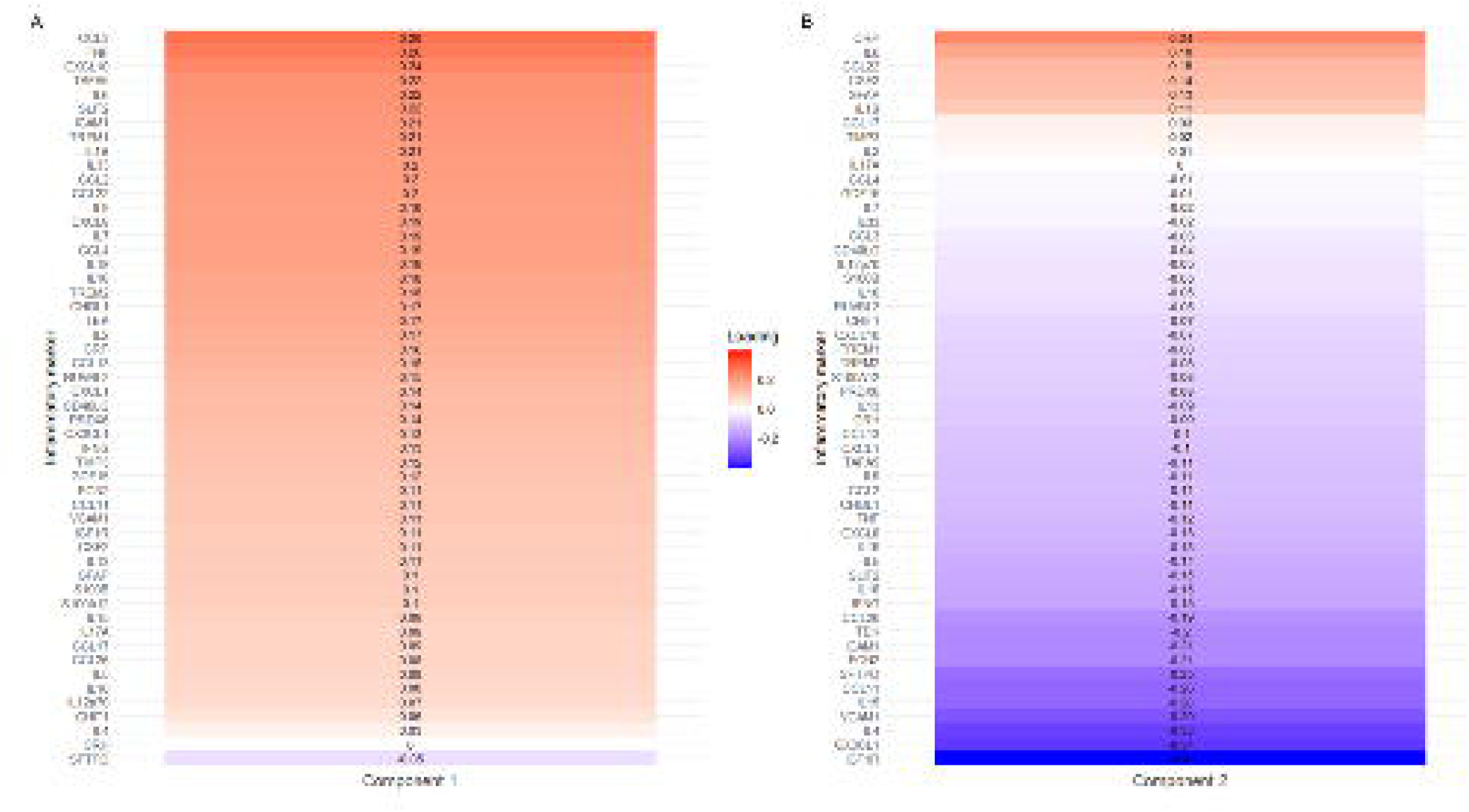
Loadings of PLS component 1 (A) and 2 (B) displaying the contribution of each inflammatory marker to the respective components. Colours represent strength and direction of loadings.

Higher scores for PLS component 2 were also observed in FTLD patients compared with controls, t (59.78) = 2.87, p = 0.006 (Figure 4A). Significant subgroup differences were present, F (2, 62) = 5.14, p = 0.009. Post hoc Tukey testing showed a significant mean difference (MD) between bvFTD patients and controls (MD = 1.55, p = 0.007), whereas no significant differences were observed for the other subgroup comparisons (bvFTD vs PSP: MD = 0.75, p = 0.36; Control vs PSP: MD = 0.80, p = 0.24) (Figure 4C).

**Figure 4.**
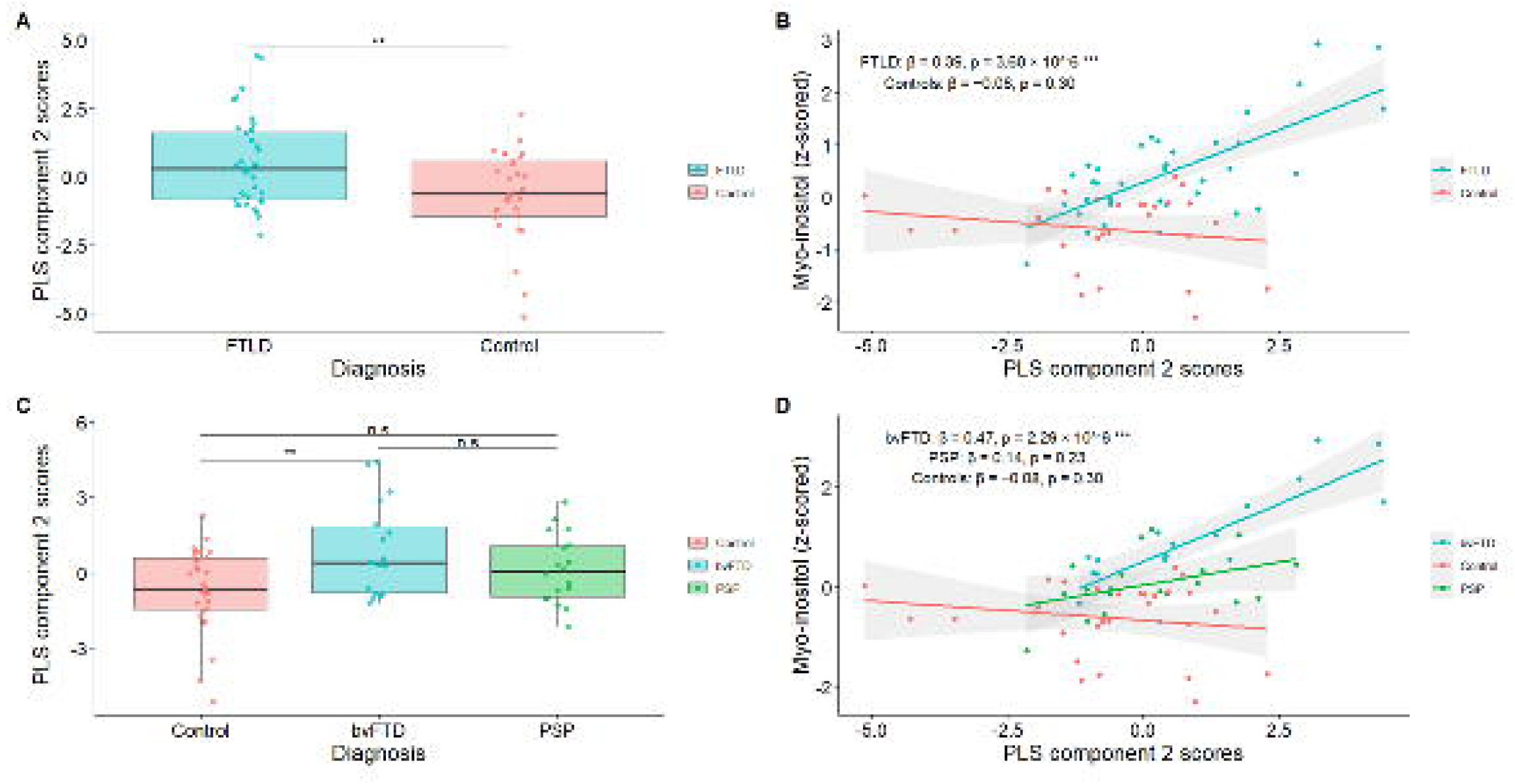
Comparison of PLS component 2 scores between FTLD participants and controls (A) and subgroups (C). Scatterplot of PLS component 2 scores against z-scored myo-inositol levels from the right inferior frontal gyms for FTLD and control groups (B) and subgroups (D). **p < 0.01 ; ***p < 0.001; n.s = not significant.

Component 2 scores were significantly associated with mI in FTLD patients (β = 0.39, SE = 0.08, t = 5.12, p = 3.60 × 10^-6), but not in controls (β = −0.08, SE = 0.08, t = −1.06, p = 0.30) (Figure 4B). The difference in slopes between groups was also significant (β = 0.47, SE = 0.11, t = 4.34, p = 5.74 × 10^-5). When subgroup-specific associations were examined (Figure 4D), the slope was significant in bvFTD (β = 0.47, SE = 0.09, t = 5.27, p = 2.29 × 10^-6), but not in PSP (β = 0.14, SE = 0.12, t = 1.20, p = 0.23). Comparisons of slopes between subgroups are shown in Table 2, with the greatest difference observed between bvFTD and controls. PSP patients showed an intermediate slope, although these differences did not reach statistical significance.

**Table 2.**
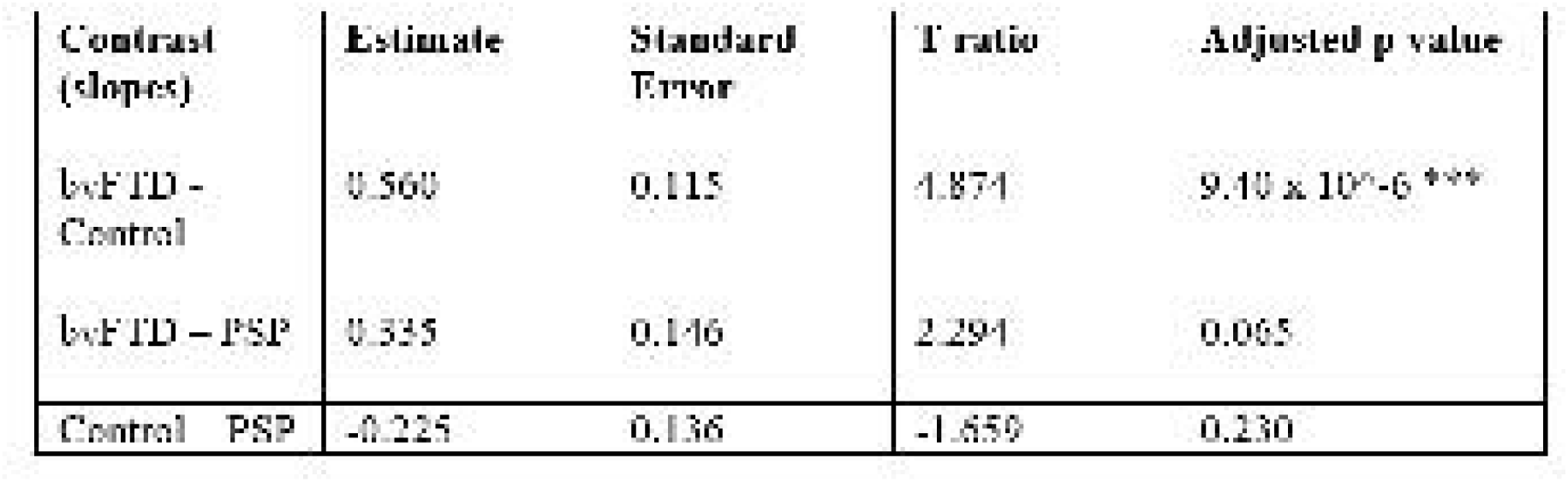
Pairwise comparisons of subgroup-specific slopes for the association between PLS component 2 and myo-inositol. ***p < 0.001.

### Associations between myo-inositol and cognition in people with bvFTD and PSP

Myo-inositol values from the RIFG were negatively associated with ACE-R score across all patients (β = −44.59, SE = 10.83, t = −4.12, p = 0.00016; Figure 5A). This negative association was present in bvFTD (β = −39.27, SE = 12.39, t = −3.17, p = 0.003), but not in PSP alone (β = −2.31, SE = 21.31, t = −0.11, p = 0.91; Figure 5B). Given that PET signals of neuroinflammation were negatively associated with the attention and orientation subscore^14^, we also examined this subscore separately: the negative association between mI and the attention and orientation subscore was observed across all patients (β = −11.33, SE = 2.46, t = −4.62, p = 3.27 × 10^-5; Figure 5C), in bvFTD (β = −11.07, SE = 2.89, t = −3.83, p = 0.0004), but not in PSP alone (β = −1.52, SE = 4.97, t = −0.31, p = 0.76; Figure 5D).

**Figure 5.**
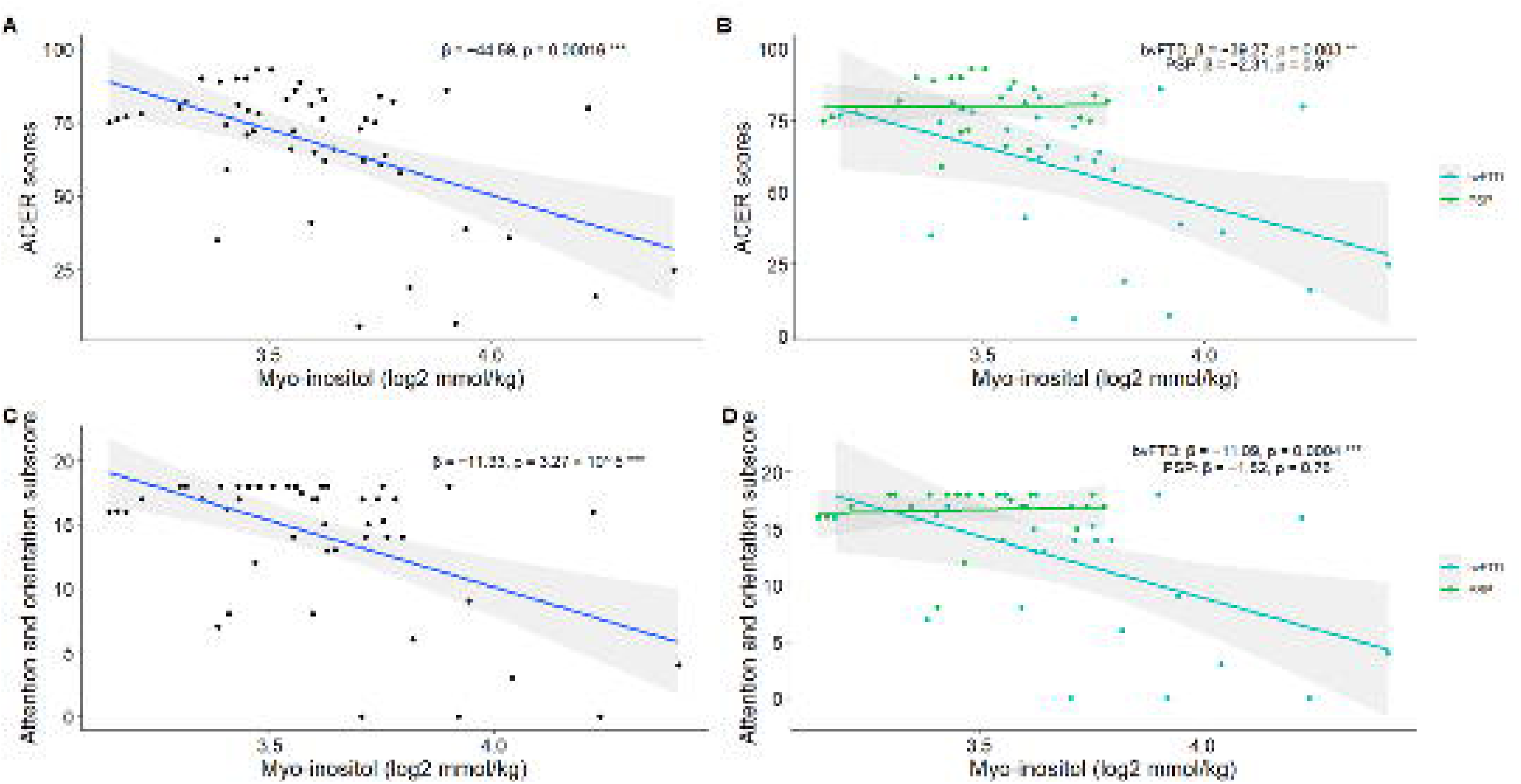
Scatterplot of 1nyo-inositol levels from the R IFG (log2 transformed) against ACER scores for the vvhole FTLD group (A), and bvFTD and PSP subgroups (B). Scatterplot of 1nyo-inositol levels from the R IFG (log2 transformed) against ACER attention and orientation subscores for all FTLD participants (C) and bvFTD and PSP subgroups (D). ***p < 0.001; **p < 0.01

## Discussion

The main finding of this study is to confirm that prefrontal myo-inositol is selectively elevated in people with FTLD; and correlates with both peripheral blood immune markers and cognitive severity. The corresponding peripheral inflammatory profile was characterised by elevated IL-6, IL-1β, GFAP, CSF2, CCL22, and CRP. These effects were not observed in the occipital control region, nor in progressive supranuclear palsy during subgroup analysis.

Our findings confirm the elevation of myo-inositol in frontotemporal dementia^24–26^. The difference from previous negative reports may be due to the greater sensitivity and specificity of MRS at 7T versus 3T. Our cohort also included people with PSP, for whom the difference in myo-inositol was not significant but was in the same direction as bvFTD. Although the prefrontal cortex is associated with significant cortical pathology in PSP, this may not have been sufficient to be detected in our cohort using the RIFG voxel. Subcortical MRS may show elevated myo-inositol but lies outside the data available from this study.

We confirm the proposed association between myo-inositol and the blood biomarkers of inflammation. The composition of this profile (the PLS latent variable 2) supports the interpretation of myo-inositol as an *in vivo* marker of neuroinflammation, reflecting an inflammatory state with astrocytic and microglial processes (IL-1β, IL-6, GFAP, CSF2, CCL22, CRP). Of the bioanalytes weighted positively in the latent inflammatory variable, IL-1β is a member of the interleukin-1 cytokine family and a key mediator of inflammatory responses including proliferation, differentiation, and apoptosis^48,49^. IL-6 is a pleiotropic cytokine that coordinates innate and adaptive immune responses and contributes to both the acute and chronic phases of inflammation^50,51^. Both are elevated in blood in bvFTD and PSP^52,53^. GFAP is primarily an astrocytic marker^54^, with post-mortem examinations of FTD and PSP patients showing elevated cortical GFAP expression^55^ while serum GFAP is increased across the FTLD spectrum^56^. This also mirrors findings in Alzheimer’s disease where GFAP was correlated with myo-inositol^57^. CSF2 regulates the production, differentiation and function of granulocytes, macrophages and is a strong microglial mitogen^58–61^. CCL22 displays chemotactic activity for monocytes, dendritic cells, and natural killer cells acting via the CCR4 receptor^62^. Elevated levels of CCL22 have also been noted in genetic frontotemporal dementia^63^. Moreover, CCL22 can be produced by microglia and astrocytes, with CCR4 expression reported in both cell types^64,65^. 7T MR spectroscopy of the brainstem following hospitalisation for COVID19 found a similar association between myo-inositol and CRP, and with disease severity^29^.

Several analytes load negatively onto PLS latent variable 2, including Surfactant Protein D (SFTPD), eotaxin-1 (CCL11), Interleukin 15 (IL-15), Vascular Cell Adhesion Molecule 1 (VCAM1), Interleukin 4 (IL-4), fractalkine (CX3CL1) and Insulin-like Growth Factor 1 Receptor (IGF1R). These negatively weighted analytes suggest that the myo-inositol-associated inflammatory profile does not represent a non-specific elevation of all immune markers. Indeed, some of the reduced biomarkers, like IL-4, are associated with anti-inflammatory and reparative microglial phenotypes^66,67^. Others, like CX3CL1 influence neuron–microglia communication pathways with context-dependent neuroprotective and immunomodulatory effects^68,69^. IGF1R signalling is also implicated in neurotrophic, metabolic and neuroinflammatory regulation, although its effects appear context dependent across ageing and neurodegeneration^70^. However, not all negatively weighted biomarkers are readily interpreted as compensatory or anti-inflammatory. CCL11 has been linked to ageing and reduced neurogenesis^71,72^, while VCAM1 is a marker of endothelial activation that has been implicated in blood–brain barrier and neuroinflammatory mechanisms^73,74^. IL-15 is also a pleiotropic cytokine involved in both protective and inflammatory immune responses, particularly through effects on NK cells and T-cell populations^75,76^. Therefore, the negative pole of this component represents a mixed peripheral immune/endothelial axis, that is arguably distinct from the glial/acute-phase inflammatory profile captured by the positive loadings.

The clinical relevance of the elevated myo-inositol is reflected in the inverse correlation with cognition. This mirrors results from TSPO PET studies of microglial activation in frontotemporal dementia^14^. However, unlike the longitudinal study of Malpetti et al., the present study was restricted to cross-sectional data only. Nonetheless, our study parallels findings identifying the impact of myo-inositol elevations on cognition in ageing^77^, the Alzheimer’s disease continuum^78,79^ and Down syndrome which is a risk factor for Alzheimer’s^80^. The associations of myo-inositol with peripheral markers and cognition were strongest in the bvFTD group, whereas the PSP cohort showed intermediate effects that did not reach statistical significance. There are several reasons for this. Although PSP is associated with impaired function of the prefrontal cortex, cognitively and physiologically^32,36,81–83^, the pathological severity is less in the RIFG than subcortical structures which might also be the case for myo-inositol. Second, the peripheral inflammatory signals in PSP may be distinct or weaker than bvFTD, weakening the correlation with myo-inositol. Similarly, there is less variance in ACE-R in our PSP cohort, reducing the power to detect correlations between cognition and myo-inositol.

There are limitations to this study. First, the analysis was retrospective and cross-sectional, with temporal differences between imaging, serum testing, and clinical assessments. The lack of group differences in net temporal offset and the inclusion of offset as a covariate both mitigate the effect, but it cannot be wholly excluded. Second, the subset of participants with both MRS and peripheral inflammatory marker data was smaller than the MRS sample. Although this impacted our power to detect more modest effects, a significant relationship with biological specificity was still identified. Third, our spectroscopic examination is limited to two voxels in specific regions rather than the whole brain. New methods for MRS-imaging, or whole brain multi-voxel MRS^84^ will enable future studies to examine widespread myo-inositol correlates. Although we could not adopt such a whole-brain approach, the RIFG was selected a priori on the basis of its role in both PSP and bvFTD pathophysiology, and prior GABA/Glutamate spectroscopic differences in the RIFG^32,35,36,85,86^. Its relationship with blood markers and cognition also enables comparison with prior PET findings^14,17^, thereby providing an additional biological validation of the MRS signal.

Overall, our findings support the potential of spectroscopy-derived myo-inositol as a neuroimaging biomarker of neuroinflammation by demonstrating: (1) elevated levels in a prefrontal region affected by FTLD; (2) associations between myo-inositol in diseased regions and specific serum biomarkers of neuroinflammation; and (3) associations between myo-inositol and cognitive decline. These observations accord with TSPO PET studies of neuroinflammation in bvFTD and PSP, using a repeatable, non-invasive method without radiation exposure. Future studies linking MRS-myo-inositol to PET, CSF and postmortem findings will further support MRS as a neuroimaging biomarker of inflammation.

## Data availability

Anonymized processed data can be shared upon request with the corresponding author. Raw data may also be requested but are likely to be subject to a data transfer agreement with restrictions required to comply with participant consent and data protection regulations.

## Acknowledgements

The authors thank the study participants and their families and carers, the radiographers at the Wolfson Brain Imaging Centre, University of Cambridge and the Centre for Magnetic Resonance Research, University of Minnesota for providing the 7T sLASER pulse sequence. The views expressed are those of the authors and not necessarily those of the NIHR or the Department of Health and Social Care. For the purpose of open access, the authors have applied a CC BY public copyright licence to any Author Accepted Manuscript version arising from this submission.

