## Supplementary Figure 1 for "Myo-inositol as a spectroscopic marker of neuroinflammation in frontotemporal lobar degeneration"

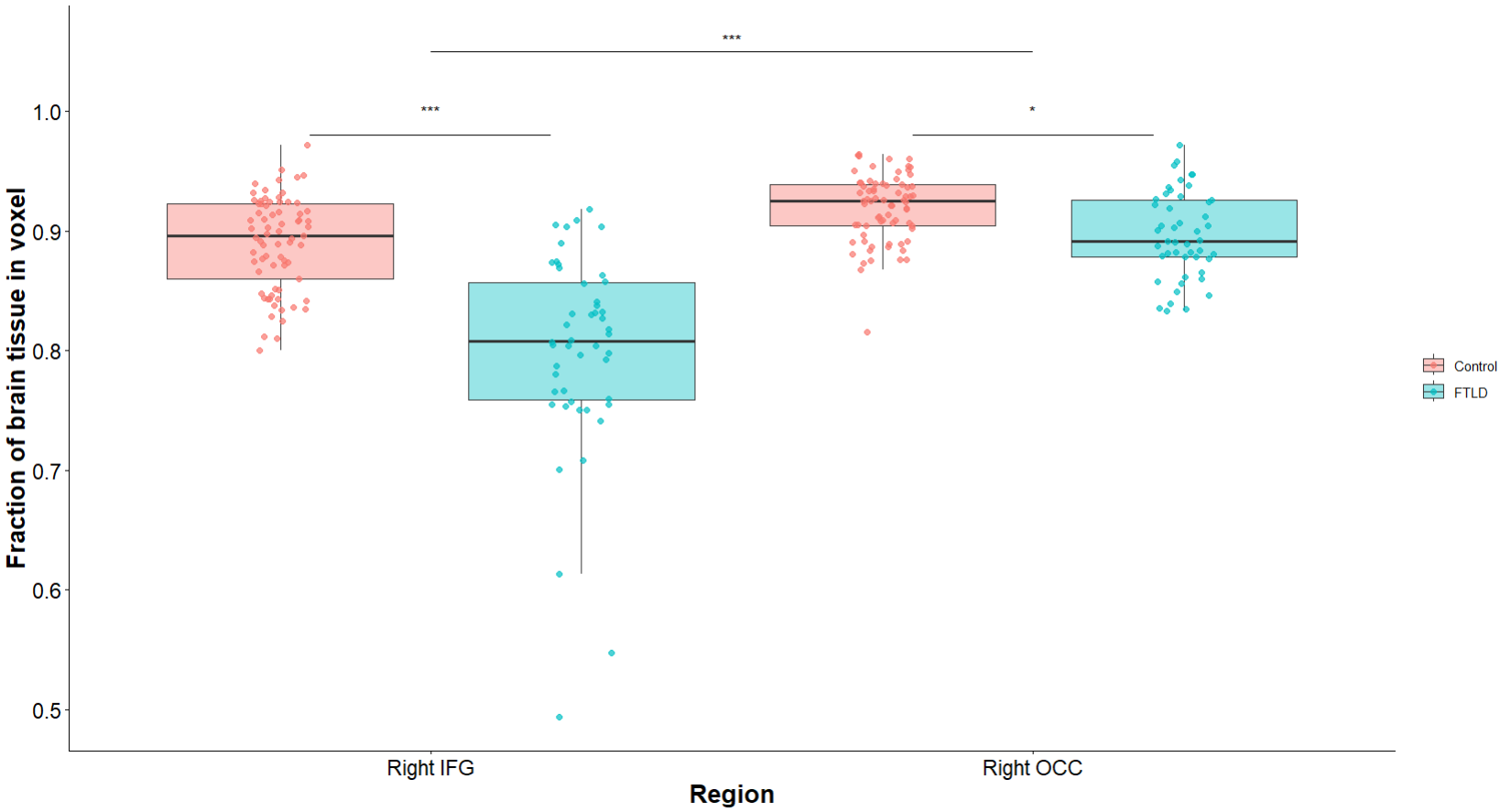

Supplementary Figure 1. Boxplots of fraction of brain tissue in voxels (1 – CSF) for voxels in the right inferior frontal gyrus (IFG) and right occipital lobe (OCC) by FTLD and control groups. *** p < 0.001; * p < 0.05.
