## Supplementary Figure 2 for "Myo-inositol as a spectroscopic marker of neuroinflammation in frontotemporal lobar degeneration"

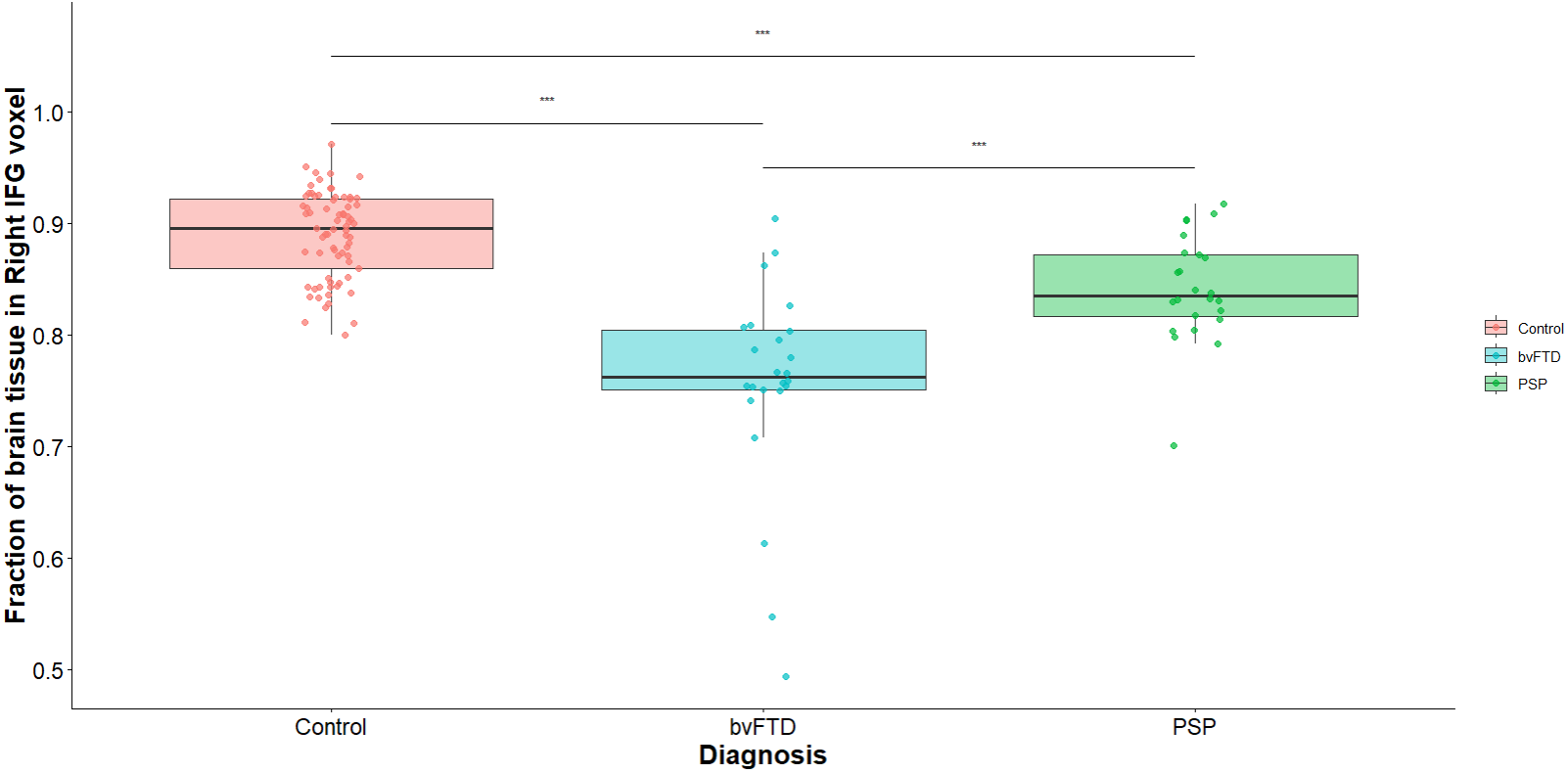

Supplementary Figure 2. Boxplots of fraction of brain tissue in voxels (1 – CSF) for voxels in the right inferior frontal gyrus (IFG) by subgroups. *** p < 0.001
