## Supplementary Figure 3 for "Myo-inositol as a spectroscopic marker of neuroinflammation in frontotemporal lobar degeneration"

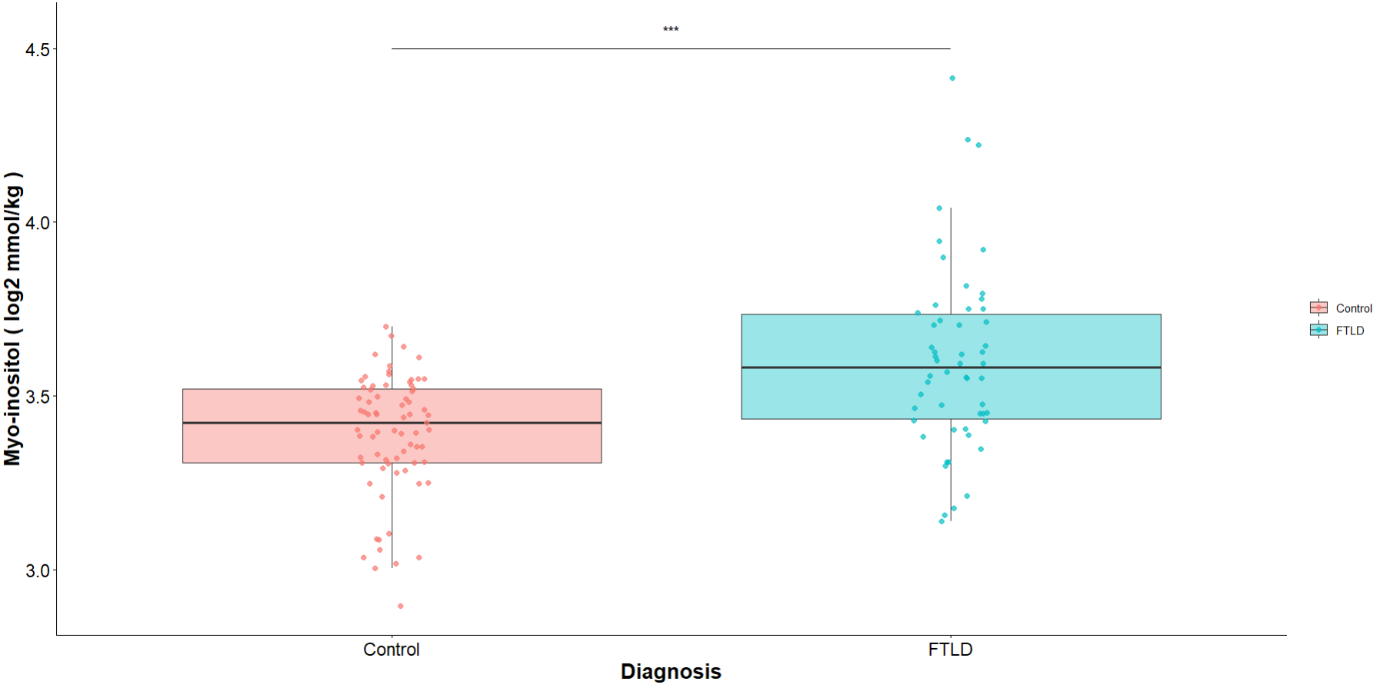

Supplementary Figure 3. Boxplots of myo-inositol levels for FTLD and control groups in the right inferior frontal gyrus. *** p < 0.001
