## Supplementary Figure 4 for "Myo-inositol as a spectroscopic marker of neuroinflammation in frontotemporal lobar degeneration"

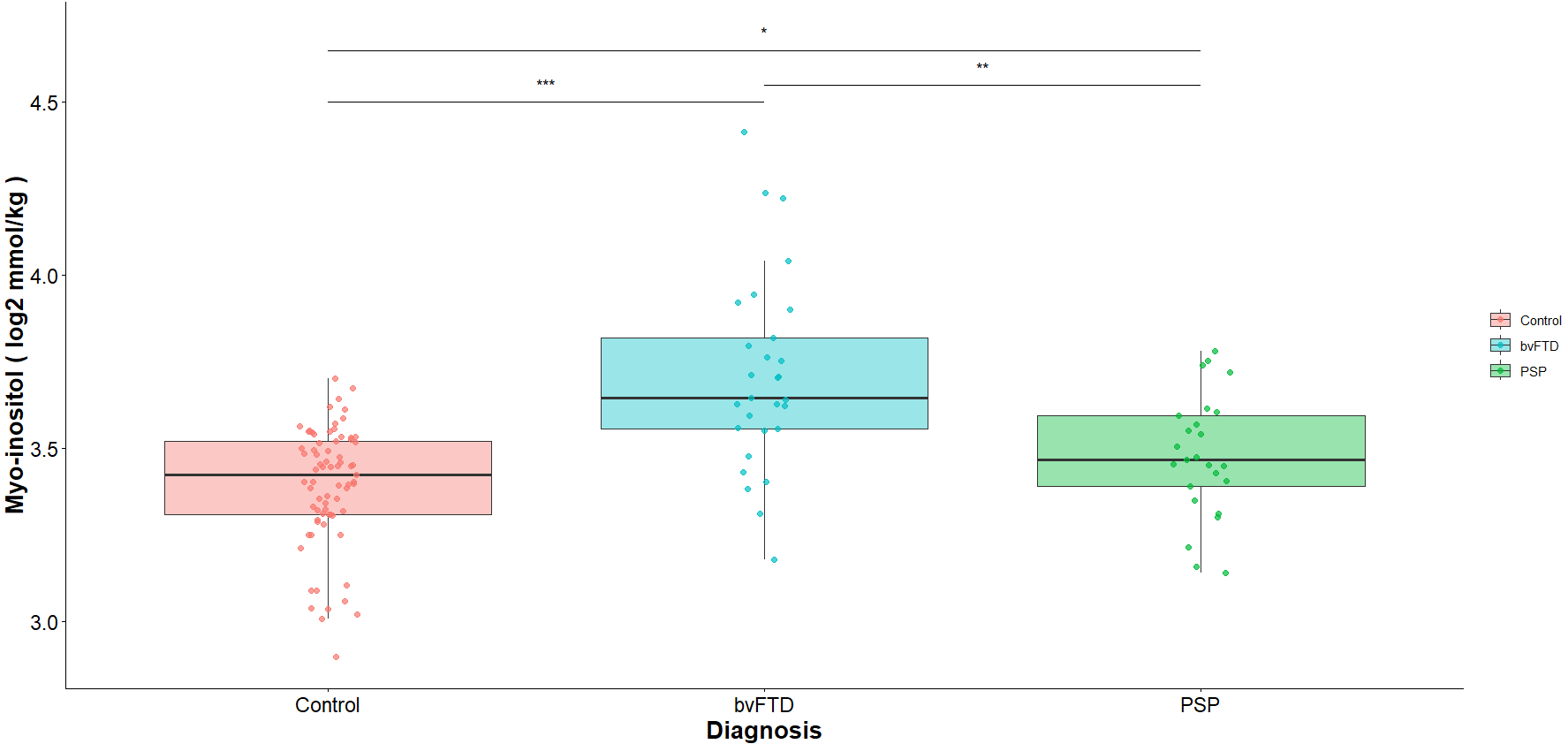

Supplementary Figure 4. Boxplots of myo-inositol levels for bvFTD, PSP and control groups in the right inferior frontal gyrus. *** p < 0.001; ** p < 0.01; * p < 0.05
