## Supplementary Figure 6 for "Myo-inositol as a spectroscopic marker of neuroinflammation in frontotemporal lobar degeneration"

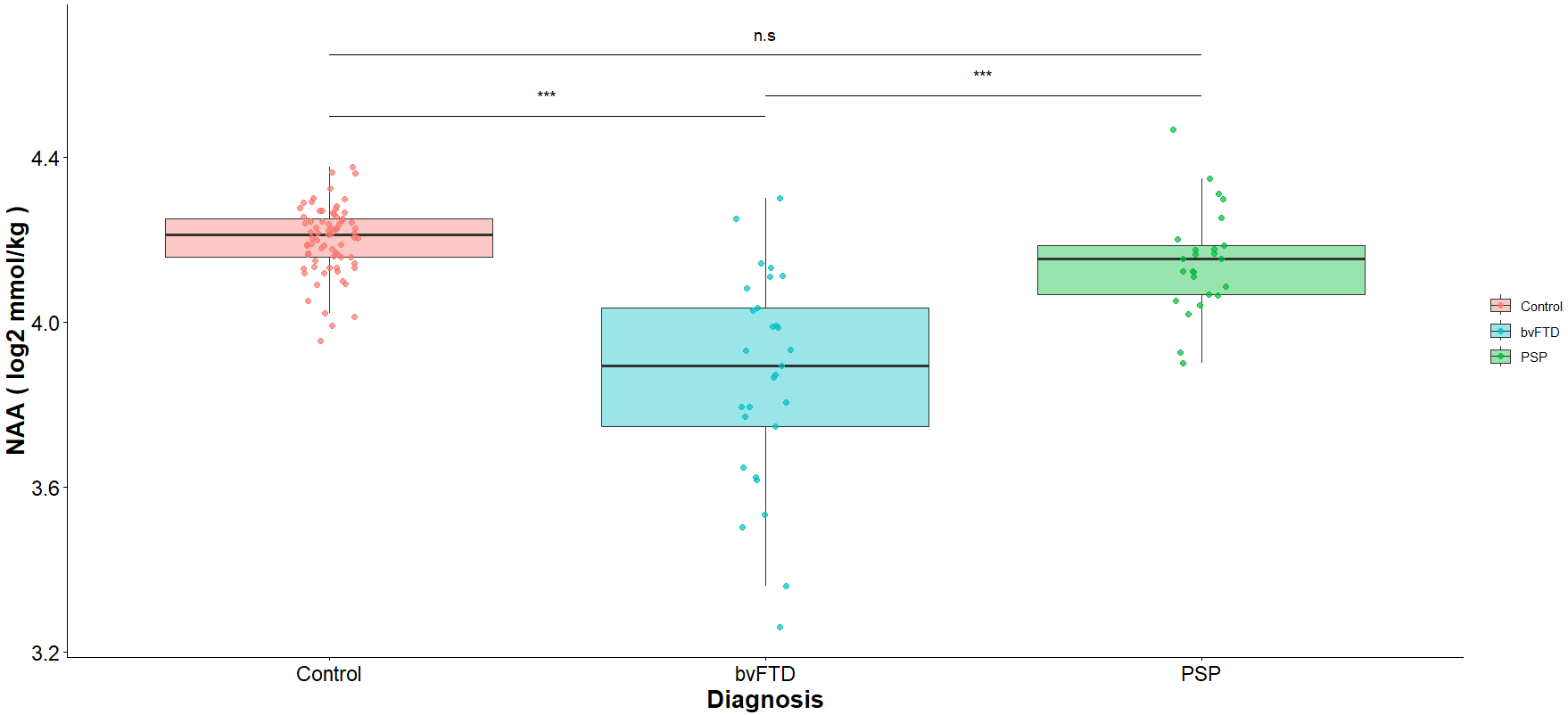

Supplementary Figure 6. Boxplots of NAA levels for bvFTD, PSP and control groups in the right inferior frontal gyrus. *** p < 0.001; n.s = not significant
