## Supplementary Table 1 for "Myo-inositol as a spectroscopic marker of neuroinflammation in frontotemporal lobar degeneration"

| **Metabolite** | **Group** | **SNR** | **Stats** | **FWHM** | **Stats** | **CRLB** | **Stats** |
| --- | --- | --- | --- | --- | --- | --- | --- |
| Myoinositol (R IFG) | Controls | 43.32 +/- 17.31 | Controls vs FTLD t = 1.60, p = 0.113 | 43.92 +/- 3.06 | Controls vs FTLD t = -3.11, p = 0.002 | 0.23+/- 0.03 | Controls vs FTLD t = -2.56, p = 0.013 |
|  | FTLD | 39.66 +/- 7.89 |  | 45.23+/- 1.62 |  | 0.26 +/- 0.09 |  |

Supplementary Table 1. Comparison of MRS quality-control metrics for myo-inositol between control and FTLD participants in the right inferior frontal gyrus (R IFG). Metrics included signal-to-noise ratio (SNR), full width at half maximum (FWHM), and Cramér–Rao lower bounds (CRLB). Values are presented as mean ± standard deviation.
