## Supplementary Table 2 for "Myo-inositol as a spectroscopic marker of neuroinflammation in frontotemporal lobar degeneration"

| **Metabolite** | **Group** | **SNR** | **Stats** | **FWHM** | **Stats** | **CRLB** | **Stats** |
| --- | --- | --- | --- | --- | --- | --- | --- |
| Myoinositol (R OCC) | Control | 48.70 +/- 15.50 | HC vs FTLD t = 2.12, p = 0.037 | 45.03+/- 1.15 | HC vs FTLD t = -0.65, p = 0.52 | 0.23+/- 0.08 | HC vs FTLD t = 2.32, p = 0.02 |
|  | FTLD | 42.60 +/- 15.32 |  | 45.17 +/- 1.08 |  | 0.21 +/- 0.04 |  |

Supplementary Table 2. Comparison of MRS quality-control metrics for myo-inositol between control and FTLD participants in the right occipital lobe (R OCC). Metrics included signal-to-noise ratio (SNR), full width at half maximum (FWHM), and Cramér–Rao lower bounds (CRLB). Values are presented as mean ± standard deviation.
