## Supplementary Table 3 for "Myo-inositol as a spectroscopic marker of neuroinflammation in frontotemporal lobar degeneration"

| ACHE | CD63 | GFAP | IL6R | PDLIM5 | SMOC1 |
| --- | --- | --- | --- | --- | --- |
| AGRN | CHI3L1 | GOT1 | IL7 | PGF | SNAP25 |
| ANXA5 | CHIT1 | HBA1 | IL9 | PGK1 | SNCA |
| APOE | CNTN2 | HTT | KDR | POSTN | SNCB |
| ARSA | CRH | ICAM1 | KLK6 | PRDX6 | SOD1 |
| Aβ38 | CRP | IFNG | MAPT | PSEN1 | SQSTM1 |
| Aβ40 | CSF2 | IGF1R | MDH1 | pSNCA-129 | TAFA5 |
| Aβ42 | CST3 | IGFBP7 | MME | p-tau-181 | TARDBP |
| BACE1 | CX3CL1 | IL10 | MSLN | p-tau-217 | TEK |
| BASP1 | CXCL1 | IL12p70 | NEFH | p-tau-231 | TIMP3 |
| BDNF | CXCL10 | IL13 | NEFL (or NfL) | pTDP43-409 | TNF |
| CALB2 | CXCL8 | IL15 | NGF | PTN | TREM1 |
| CCL11 | ENO2 | IL16 | NPTX1 | REST | TREM2 |
| CCL13 | FABP3 | IL17A | NPTX2 | RUVBL2 | UCHL1 |
| CCL17 | FCN2 | IL18 | NPTXR | S100A12 | VCAM1 |
| CCL2 | FGF2 | IL1B | NPY | S100B | VEGFA |
| CCL22 | FLT1 | IL2 | NRGN | SAA1 | VEGFD |
| CCL26 | FOLR1 | IL33 | Oligo-SNCA | SFRP1 | VGF |
| CCL3 | GDF15 | IL4 | PARK7 | SFTPD | VSNL1 |
| CCL4 | GDI1 | IL5 | PDGFRB | SLIT2 | YWHAZ |
| CD40LG | GDNF | IL6 |  |  |  |

Supplementary Table 3. Full list of List of targets in NULISAseq CNS panel.
