## Supplementary Table 4 for "Myo-inositol as a spectroscopic marker of neuroinflammation in frontotemporal lobar degeneration"

| CCL11 | CHI3L1 | FCN2 | IL15 | IL6 | SLIT2 |
| --- | --- | --- | --- | --- | --- |
| CCL13 | CHIT1 | GDF15 | IL16 | IL7 | TAFA5 |
| CCL17 | CRH | GFAP | IL17A | IL9 | TEK |
| CCL2 | CRP | ICAM1 | IL18 | PRDX6 | TIMP3 |
| CCL22 | CSF2 | IFNG | IL1B | RUVBL2 | TNF |
| CCL26 | CX3CL1 | IGF1R | IL2 | S100A12 | TREM1 |
| CCL3 | CXCL1 | IL10 | IL33 | S100B | TREM2 |
| CCL4 | CXCL10 | IL12p70 | IL4 | SFTPD | VCAM1 |
| CD40LG | CXCL8 | IL13 | IL5 |  |  |

Supplementary Table 4. List of inflammatory targets in NULISAseq CNS panel.
