## Supplementary Table 5 for "Myo-inositol as a spectroscopic marker of neuroinflammation in frontotemporal lobar degeneration"

| **Region** | **Contrast (myo-inositol levels)** | **Estimate** | **Standard Error** | **T ratio** | **Adjusted p value** |
| --- | --- | --- | --- | --- | --- |
| **R IFG** | bvFTD - Control | 0.316 | 0.043 | 7.312 | 3.03 x 10^^-11^ |
|  | bvFTD – PSP | 0.199 | 0.054 | 3.690 | 0.001 |
|  | Control – PSP | -0.116 | 0.046 | -2.528 | 0.034 |

Supplementary Table 5. Subgroup comparisons of adjusted mean differences in myo-inositol levels in the right inferior frontal gyrus
