## Supplementary Table 6 for "Myo-inositol as a spectroscopic marker of neuroinflammation in frontotemporal lobar degeneration"

| **Region** | **Contrast (NAA levels)** | **Estimate** | **Standard Error** | **T ratio** | **Adjusted p value** |
| --- | --- | --- | --- | --- | --- |
| **R IFG** | bvFTD - Control | -0.335 | 0.033 | -10.163 | p < 2 X 10^-16^ |
|  | bvFTD – PSP | -0.287 | 0.041 | -6.963 | 1.81 x 10^-10^ |
|  | Control – PSP | 0.048 | 0.035 | 1.361 | 0.364 |

Supplementary Table 6. Subgroup comparisons of adjusted mean differences in NAA levels in the right inferior frontal gyrus
